# Current state of prevention of mother-to-child transmission of HIV in informal health centers in Douala and Ebolowa, Cameroon

**DOI:** 10.1101/2022.08.03.22278376

**Authors:** Lydie Audrey Amboua-Schouamé, Jean Joel Bigna, Isacar Lucel Schouamé, Sylvie Kwedi Nolna, Antoine Socpa

**Author notes:** Corresponding author: Lydie Audrey Amboua-Schouamé, (ASLA).

## Abstract

**Background:** Informal health care providers are key actors for health care provision in developing countries for poor populations. Thus, in Cameroon, in 2017, there were more than 3,000 Informal Health facilities. In a context of elimination of mother-to-child transmission of HIV, we describe the offer of Prevention of Mother to Child Transmission of HIV (PMTCT) in informal health centers.

**Methods:** This two-phase cross-sectional study was carried out in two cities in Cameroon notably Douala and Ebolowa. The first step was conducted from March 2019 to July 2019 in 110 informal health centers and the second from August 2019 to January 2020 with 183 Health Providers in these informal health centers. Standardized questionnaires were administered. Data collected were entered in kobo Collect software. Descriptive statistics and logistic regression at a level of significance of 5% were used.

**Results:** Of the 110 informal health centers, 109 integrated HIV testing into the antenatal check-up package. Among them, 43% (47/109) reported giving antiretroviral treatment to HIV infected pregnant women, while the remaining referred them to formal HIV care centers. Regarding delivery services, 52% (53/101) of those who offered them referred HIV exposed newborn for further PMTCT care. Knowledge of PMTCT was insufficient among 51% (94/183) of health providers and 90% (165/183) had insufficient PMTCT practices. Health providers with no PMTCT experience were more likely to have insufficient knowledge of PMTCT (aOR= 32.04, 95%CI: 6.29 to 163.10, p<0.001) whereas, those without any formal PMTCT training were more at risk of having insufficient knowledge (aOR= 3.02, 95%CI: 1.06 to 8.64, p=0.03) and insufficient practices (OR= 4.35, 95%CI: 1.44 to 13.09, p=0.009) towards PMTCT.

**Conclusion:** Given their proximity to the populations and the PMTCT activities they conduct most often; particular attention should be paid to PMTCT in informal health centers.

## Introduction

The various economic crises that have hit African countries as well as the following structural adjustment policies from mid-80s had a serious impact on the creation of jobs and services **[1,2]**. Consequently, an exponential increase of populations impoverishment and informal activities were observed **[2]**. The latter will develop and spread in all sectors of activity, including health sector. Following the impoverishment of populations in low-income countries, they mainly resort to the informal health system for their health cases management. This massive recourse was explained by its geographical proximity, its financial accessibility and the flexibility of the mode of payment which is practiced there, as well as the reduced waiting times **[3–6]**.

In Cameroon, the economic crisis of the 1980s led to the reduction of public health expenditure. These decrease from about 5% of the national budget in 1994 to less than 3% in the following years **[7]**. The immediate consequence was the deterioration of the health system and the quality of care and the parallel exponential rise of a vast network of informal health providers **[8–11]**. This huge informal health network includes several actors: street medicine vendors, traditional healers and Informal Health Centers (IHCs). In 2017, the Ministry of Public Health made a census of 3,058 informal health facilities operating on the national territory**[12]**.They operate without an official act of creation or operation issued by the government. However, they are subject to taxation which seems paradoxical.

These IHCs, despite their limited technical facilities, generally provide a wide range of services, integrating antenatal consultations and childbirths services **[13,14]**. Antenatal consultations (ANC) are an essential aspect for the prevention of HIV transmission from mother to child (PMTCT), since it represents a critical entry point into the PMTCT cascade **[15,16]**. Especially in our context, client demand-based HIV testing is very low. Thus, the ANC remains the major method for HIV testing in women **[17]**. Moreover, ANC is all the more important since the majority of pediatric HIV cases result from mother-to-child transmission **[18–23]**.

Currently, nothing is known about PMTCT activities concerning the ICHs in Cameroon. As part of the ECIP PMTCT study, carried out in the cities of Douala and Ebolowa, we evaluated PMTCT in informal health centers. This paper aims to present the current state of PMTCT in IHCs in these two cities. Specifically, this work involved analyzing the PMTCT offer and assessing knowledge and practices of PMTCT. We also determined factors associated with the “insufficient” knowledge and practices among Healthcare Providers (HP) in the IHCs.

## Materials and methods

### Study design setting and population

We conducted a two-phases descriptive cross-sectional study in Douala and Ebolowa, two cities in Cameroon. The first phase was carried out from March to July 2019 and involved IHCs offering antenatal care and or childbirth service. The second phase, carried out from August 2019 to January 2020 involved HPs working in the above-mentioned services. A total of 110 IHCs were registered including 90 in Douala and 23 in Ebolowa; while 183 HPs were enrolled including 131 in Doula and 52 in Ebolowa.

### Procedure

#### Definition of Informal Health Centers

We define an informal health center as any secular private health facility with a fixed location, without an official, government-issued, act of creation or operation, and without physicians.

#### Selection of informal health centers

To select the IHCs, we used the health map of the city of Douala and Ebolowa. Cameroon’s health map divides each city into health districts. In the city of Douala, we have 9 Health Districts: Boko, Japoma, Nylon, Bangue, Deido, Cité des Palmiers, Bonassama, Newbell and Logbaba. Health Districts were randomized using the Excel spreadsheet. This randomization allowed us to retain 7 health districts: Nylon, Bangue, Deido, Cité des Palmiers, Bonassama, Newbell and Logbaba. Once the Health Districts were known in the two research locations, field workers recruited for the study had a starting point in each Health District. This starting point was the benchmark for combing the informal health centers within the entire Health District Since the city of Ebolowa has only one Health District, the entire district was retained. An inventory of IHCs operating in Ebolowa was generated and a number of them were selected for inclusion in the study through a random process.

### Data collection and Statistical analysis

Based on PMTCT assessment tool developed by Family Health international **[24]**, we developed a questionnaire to capture information relating to PMTCT offer. The questionnaire collected information on availability of ANC services, integration of HIV testing in ANC package, supply of Antiretroviral Treatment (ART) to HIV Infected Pregnant Women (HIVPW), ART supply source, availability of delivery services, availability of ARV for HIV exposed newborn, HIV tests used to incriminate an HIV infection, duration of the mother-child couple follow-up for IHCs providing on-site HIV care.

Furthermore, Health staff were interviewed using standardized questionnaires to assess their PMTCT Knowledge and Practices. The questionnaire was designed from the national guidelines for prevention and care of HIV in Cameroon **[25]**. Overall 14 and 7 questions evaluating PMTCT knowledge and practices, respectively, were asked to HPs.

The questionnaire also has an important section on Sociodemographic data (town, type of PMTCT site, gender, health training qualification, type of PMTCT training received, years of experience in PMTCT experience, history of formal PMTCT training during the last 2 years).

Regarding PMTCT knowledge, questions asked were on : HIV source; main route of HIV infection in children; lack of ARV coverage in HIVPW and risk of HIV infection in the child; lack of ARV prophylaxis coverage in the child and risk of HIV infection; ARV protocol recommended as first line treatment for HIVPW; delay of ART initiation in HIVPW; delay of ARV prophylaxis in HIV exposed children; recommended ARV prophylaxis in exposed children; duration of this prophylaxis; knowledge of early diagnosis for HIV exposed children; recommended age for HIV early diagnosis for HIV exposed children; Cotrimoxazole initiation delay in HIV-exposed children; recommended timing for stopping Cotrimoxazole in HIV exposed children; breastfeeding for HIV exposed children.

While for PMTCT practices, the questions were on the proposal of HIV testing to pregnant women; the practice of HIV pre- and post-test counseling; the proposal/reminder of the importance of ARV treatment to HIVPW; encouragement of HIV status disclosure to male partners; administration / reminder of the administration of ARV prophylaxis to HIV exposed children; reminder to the HIV mothers of the importance of early infant diagnosis of HIV for exposed children; reference procedure for further PMTCT services. All the data collected were entering in Kobo Collect.

For sociodemographic variables, descriptive analyses were performed. To assess PMTCT knowledge and practices, for each good response gave, a score of one point was assigned and zero otherwise. The total score that an HP could therefore have oscillated between 0 and 14 for PMTCT knowledge and 0 to 7 for PMTCT practices. Afterwards, we calculated each scoring category as the percentage of correct answers divided by corresponding questions answered. The overall knowledge and practices were categorized as “insufficient” if the score was less than 80% and “sufficient” if 80% or above **[26]**. Multivariate logistic regression was used to identify associated factors to insufficient knowledge and practice of PMTCT among HPs at a level of significance of 5%.

### Ethical considerations

The ECIP-PMTCT study was approved by the National Ethics Committee of Cameroon (Ethical clearance n° 2019/03/1155/CE/CNERSH/SP) and by the Cameroonian Ministry of Public Health (Administrative authorization n° 631-15.19) before the implementation of the study. Informed and signed consent of Head of IHCs and HPs were also collected.

## Results

### PMTCT service offer in the ICHs

Almost all (109/110) of the IHCs integrated the HIV screening test within their antenatal check-up. For 95% (104/109), the final decision to make a diagnosis of HIV infection was preceded by the successive incrimination of HIV infection using the Determine and OraQuick tests/strips. For the remaining IHCs only the Determine test was used for this purpose. As we can see in Figure1 below, among the 109 IHCs that integrated HIV screening in ANC, 43% (47/109) reported giving ART to HIVPW. More than half of the ICHs (26/47) followed HIVPW for a period of 2 months post-delivery. The health districts were the supply source of ART for 89% (42/47) of IHCs. For the remaining 11%, they got their ART from others IHCs. For IHCs referring HIVPW to formal HIV care centers, this reference was verbal, without any documentation. Only the name of the HIV care center was communicated to the HIVPW.

**Fig 1:**
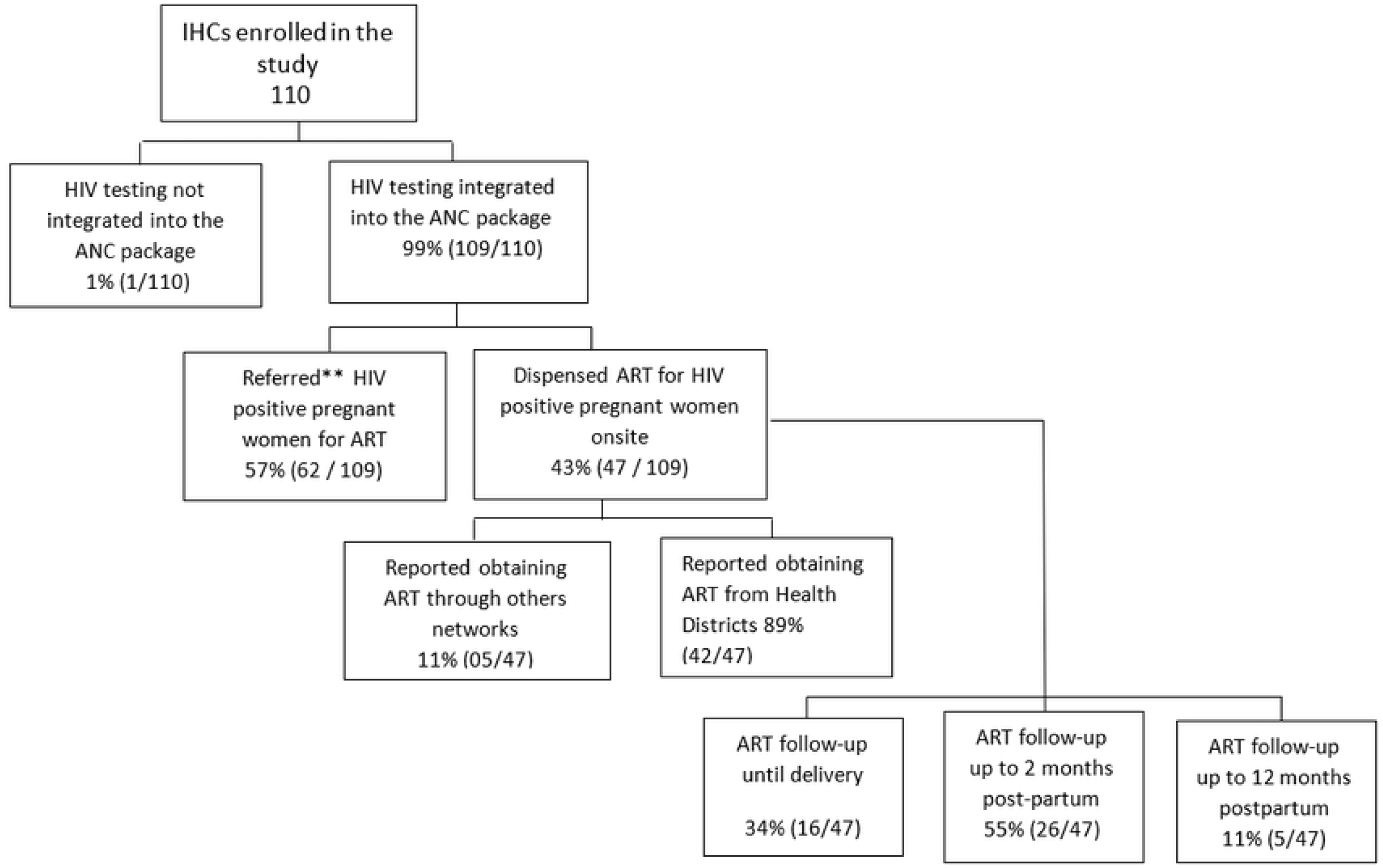
Antenatal HIV screening, ART availability and source of supply.

Regarding delivery services (Fig.2), 92% (101/110) of IHCs provided them. For 52% (53/101) of IHCs, HIV exposed children were referred for Nevirapine prophylaxis and for further PMTCT care. Meanwhile, 48% (48/101) provided Nevirapine prophylaxis to HIV exposed children. None of the IHCs dispensed Cotrimoxazole or collected blood sample for the early infant diagnosis.

**Fig 2:**
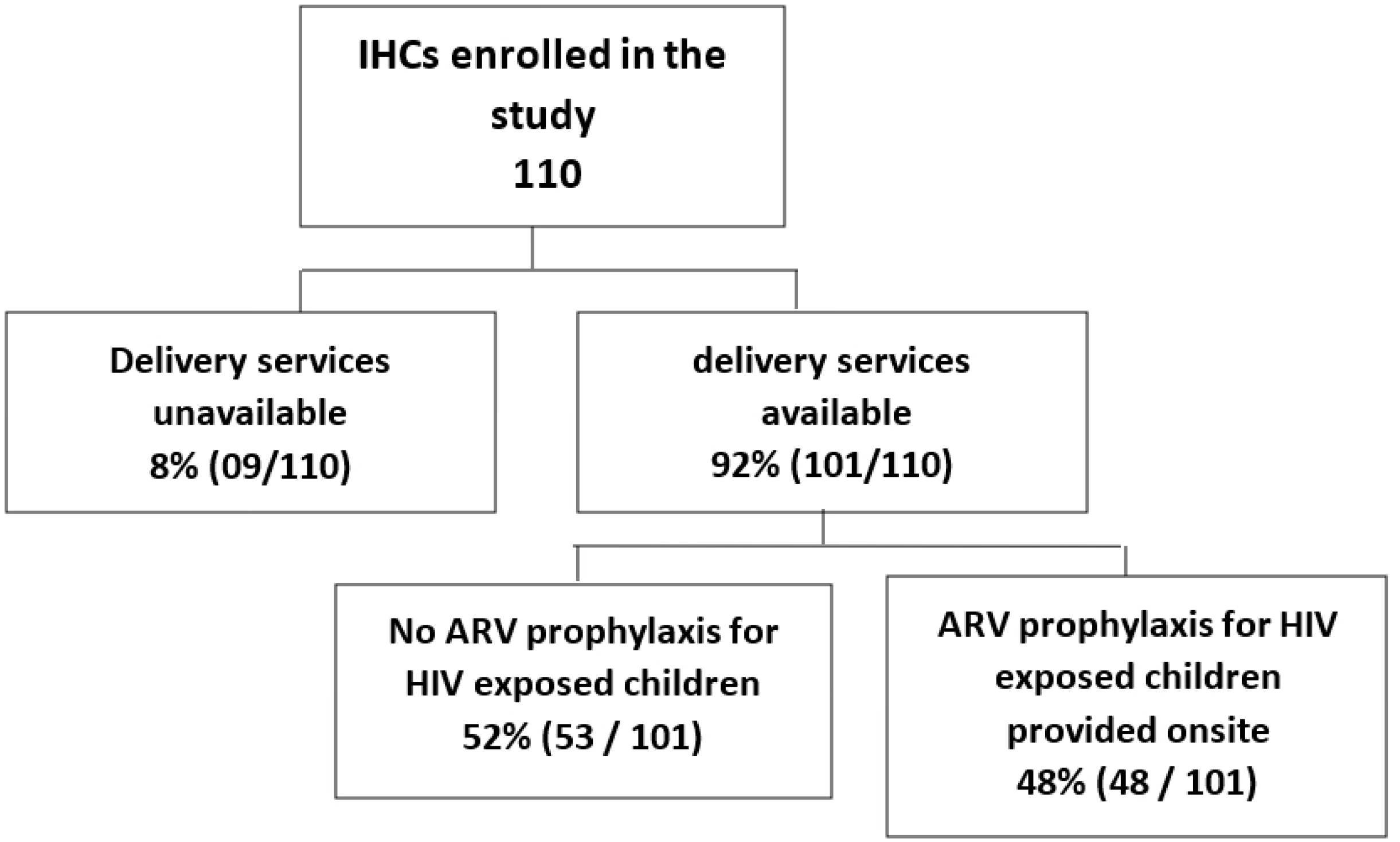
Prophylactic ARV coverage in HIV exposed children in IHCs.

### PMTCT knowledge and practices of health Providers

#### Description of HPs

A total of 183 HPs were enrolled in the study, with the median age of 37 years (IQR, 30-44). Less than a quarter (19%) were male. The majority were Nurse Assistants (56%) and we had 7% of Health staff without any formal health training. Among HPs, almost 28% (51/183) were or had been Publics Servants in public health facilities. Indeed, 84% (43/51) of them were retired and 16% (8/51) were still in activity. All those Civil Servants were owners of IHCs.

The main source of PMTCT information was colleagues (84%) whereas 13% (23/183) received a formal training through workshops or seminars during the last two years. Slightly more than half (107/183) of HPs had PMTCT experience of less than 5 years (Table 1).

**Table 1.** Characteristics of Health providers in informal health centers.

|  | Frequency | Percentage (%) |
| --- | --- | --- |
| **Town (n=183)** |  |  |
| Douala | 131 | 72 |
| Ebolowa | 52 | 28 |
| **Type of PMTCT site (n=183)** |  |  |
| Refer for PMTCT | 119 | 65 |
| PMTCT follow-up site | 64 | 35 |
| **Gender (n=183)** |  |  |
| Female | 149 | 81 |
| Male | 34 | 19 |
| **Former or current Civil Servant(n=183)** |  |  |
| Yes | 51 | 28 |
| No | 132 | 72 |
| **Health training qualification (n=183)** |  |  |
| No documented health training | 12 | 7 |
| Nurse assistant | 103 | 56 |
| State certified Nurse | 35 | 19 |
| Registered nurse | 14 | 8 |
| Laboratory technician | 19 | 10 |
| **Type of PMTCT training received (n=183)** |  |  |
| None | 2 | 1 |
| By colleagues | 154 | 84 |
| Documents reading | 4 | 2 |
| Seminars / workshops | 23 | 13 |
| **Years of experience in PMTCT(n=183)** |  |  |
| None | 25 | 14 |
| < 5 years | 107 | 59 |
| 5 – 10 years | 41 | 22 |
| >10 years | 10 | 5 |
| **Formal PMTCT training during the last 2 years (n=183)** |  |  |
| Yes | 23 | 13 |
| No | 160 | 87 |

#### PMTCT knowledge

For PMTCT knowledge, the median score was 79% (IQR, 71-93). Only 49% (89/183) of our HPs had sufficient PMTCT knowledge (scoring 80% or more). The question that registered the lowest score with 29% (58/183) was “Does an HIV infected woman on ART should breastfeed her child?” (Table.2)

**Table 2.**
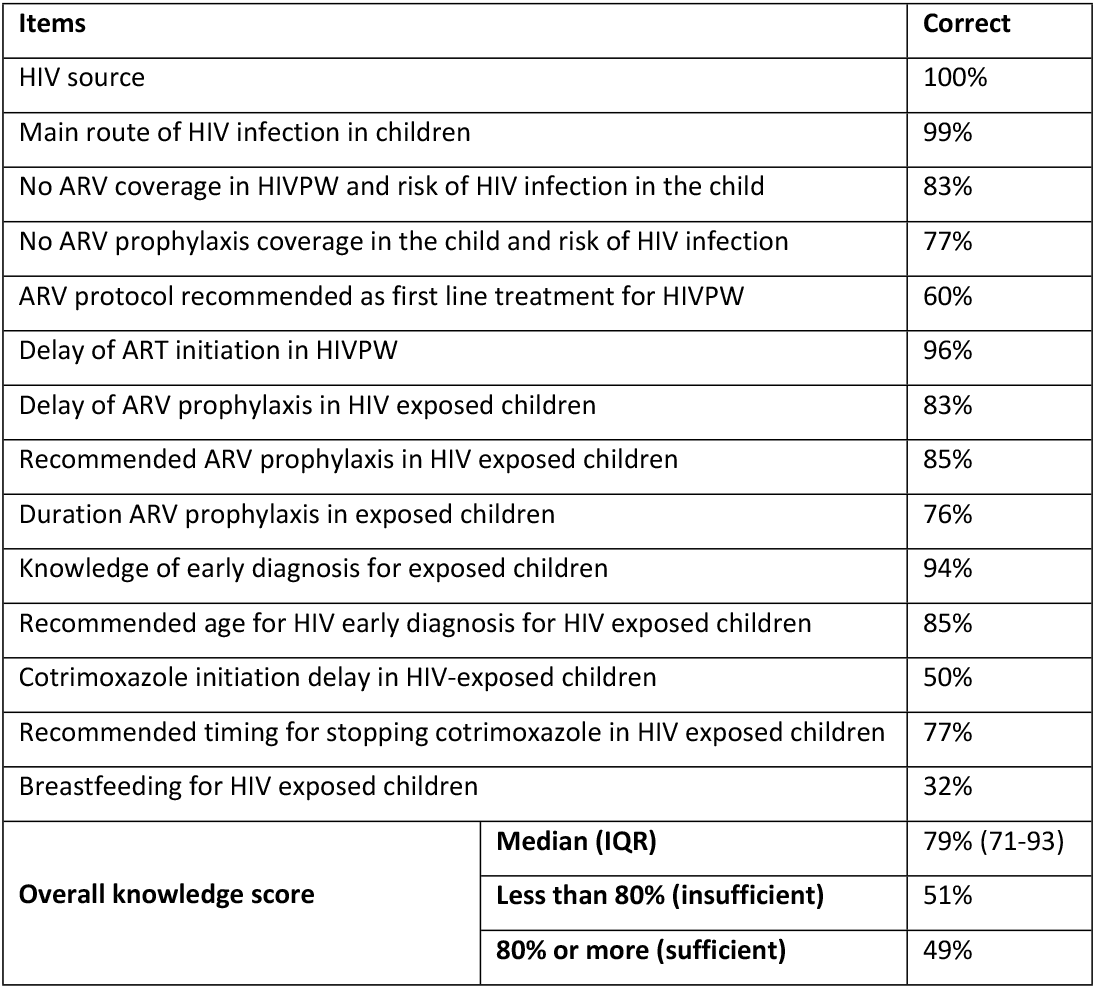
Health providers knowledge on PMTCT.

#### PMTCT Practices

Regarding PMTCT practices, the median score was 43% (IQR, 29-57). As table 3 shows, only one HP over ten (18/183) had a sufficient PMTCT knowledge. The question with the lowest score was “How do you refer an infected mother to an HIV treatment center?”, with only 8% (15/183) of good practices reported.

**Table 3.** Health Providers practices on PMTCT.

| Items |  | Correct |
| --- | --- | --- |
| Proposal of HIV testing to pregnant women |  | 100% |
| Practice of HIV pre- and post-test counseling |  | 40% |
| Proposal/reminder of the importance of ARV treatment to infected pregnant women |  | 52% |
| Encouragement of HIV status disclosure to male partners |  | 15% |
| Administration / reminder of the administration of ARV prophylaxis to exposed children |  | 52% |
| Reminder to the HIV mothers of the importance of early infant diagnosis of HIV for exposed children |  | 48% |
| Reference procedure for further PMTCT services. |  | 8% |
| Overall practice score | Median (IQR) | 43% (29-57) |
|  | Less than 80% (insufficient) | 90% |
|  | 80% or more (sufficient) | 10% |

Only about 7% (12/183) of HPs had both sufficient knowledge and practices towards PMTCT. While 48% (88/183) had neither sufficient knowledge nor sufficient PMTCT practices.

### Factors associated with poor knowledge and practices of PMTCT

In bivariate analyses, regarding health training qualification, HPs with no documented health training qualification were more likely to have insufficient PMTCT knowledge than Registered Nurses (OR= 7.35, 95% CI: 1.28 to 50, p=0.02). HPs with no PMTCT experience were forty-one time likely to have insufficient knowledge of PMTCT compared to those with 5 years or more (OR= 41.81, 95%CI: 8.51 to 205.38, p<0.001). In addition, those with less than 5 years of PMTCT experience were four time likely to do same (OR= 4.64, 95%CI: 2.15 to 10.01, p<0.001). Furthermore, HPs with no formal PMTCT training received during the past two years were slightly 3 times to have insufficient knowledge compared to those who were trained (OR= 2.72, 95%CI: 1.06 to 6.99, p=0.037).

As presented in Table 4 below, in multivariate logistic model, lack of formal PMTCT training and lack or fair experience in PMTCT were associated with insufficient knowledge of PMTCT. Indeed, HPs with no history of formal PMTCT training during the past two years were three time most at risk to have insufficient knowledge of PMTCT compared to those who were formally trained (aOR= 3.02, 95%CI: 1.06 to 8.64, p=0.03). Moreover, HPs with no PMTCT experience were thirty-two times at risk to have insufficient PMTCT Knowledge compared to those with 5 years’ experience or more (aOR= 32.04, 95%CI: 6.29 to 163.10, p<0.001). Compared to the same reference, HPs with less than 5 years of experience in PMTCT had this risk four times (aOR= 4.50, 95%CI: 2.0 to 10.15, p<0.001).

**Table 4.**
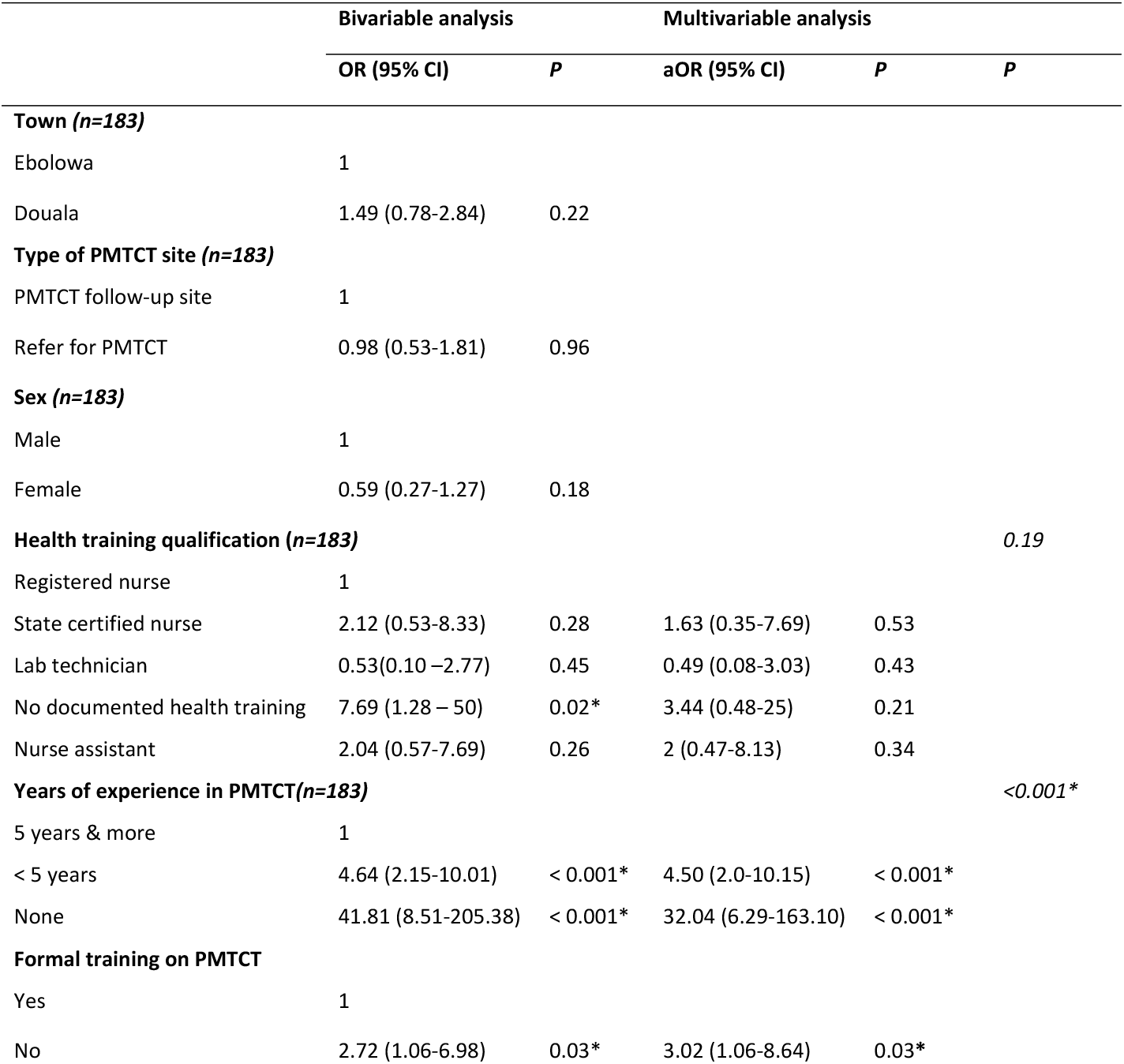
Factors associated with insufficient level of PMTCT knowledge.

|  | Bivariable analysis |  | Multivariable analysis |  |  |
| --- | --- | --- | --- | --- | --- |
|  | OR (95% CI) | P | aOR (95% CI) | P | P |
| **Town (n=183)** |  |  |  |  |  |
| Ebolowa | 1 |  |  |  |  |
| Douala | 1.49 (0.78-2.84) | 0.22 |  |  |  |
| **Type of PMTCT site (n=183)** |  |  |  |  |  |
| PMTCT follow-up site | 1 |  |  |  |  |
| Refer for PMTCT | 0.98 (0.53-1.81) | 0.96 |  |  |  |
| **Sex (n=183)** |  |  |  |  |  |
| Male | 1 |  |  |  |  |
| Female | 0.59 (0.27-1.27) | 0.18 |  |  |  |
| **Health training qualification (n=183)** |  |  |  |  | 0.19 |
| Registered nurse | 1 |  |  |  |  |
| State certified nurse | 2.12 (0.53-8.33) | 0.28 | 1.63 (0.35-7.69) | 0.53 |  |
| Lab technician | 0.53(0.10 –2.77) | 0.45 | 0.49 (0.08-3.03) | 0.43 |  |
| No documented health training | 7.69 (1.28 – 50) | 0.02* | 3.44 (0.48-25) | 0.21 |  |
| Nurse assistant | 2.04 (0.57-7.69) | 0.26 | 2 (0.47-8.13) | 0.34 |  |
| **Years of experience in PMTCT(n=183)** |  |  |  |  | <0.001* |
| 5 years & more | 1 |  |  |  |  |
| < 5 years | 4.64 (2.15-10.01) | < 0.001* | 4.50 (2.0-10.15) | < 0.001* |  |
| None | 41.81 (8.51-205.38) | < 0.001* | 32.04 (6.29-163.10) | < 0.001* |  |
| **Formal training on PMTCT** |  |  |  |  |  |
| Yes | 1 |  |  |  |  |
| No | 2.72 (1.06-6.98) | 0.03* | 3.02 (1.06-8.64) | 0.03* |  |

For insufficient PMTCT practices, HPs who worked in IHCs in Douala were more likely to have poorer practices regarding PMTCT (OR= 3.66, 95%CI: 1.35 to 9.88, p=0.01) compared to those working in Ebolowa. Furthermore, HPs who did not received any formal PMTCT training (OR= 4.35, 95%CI: 1.44 to 13.09, p=0.009) were most at risk compared to those who were formally trained. And these associations were more pronounced in multivariate logistic model as we can see in Table 5.

**Table 5.** Factors associated with insufficient level of PMTCT practices.

|  | Bivariable analysis |  | Multivariable analysis |  |  |
| --- | --- | --- | --- | --- | --- |
|  | OR (95% CI) | P | aOR (95% CI) | P | P |
| **Town (n=183)** |  |  |  |  |  |
| Ebolowa | 1 |  |  |  |  |
| Douala | 3.66 (1.35-9.88) | 0.01* | 4.09 (1.45-11.55) | **0.008\*** |  |
| **Type of PMTCT site (n=183)** |  |  |  |  |  |
| PMTCT follow-up site | 1 |  |  |  |  |
| Refer for PMTCT | 4.34 (1.54-12.21) | 0.005* |  |  |  |
| **Sex (n=183)** |  |  |  |  |  |
| Male | 1 |  |  |  |  |
| Female | 0.23 (0.03-1.83) | 0.16 |  |  |  |
| **Health training qualification (n=183)** |  |  |  |  |  |
| No documented health training | 1 |  |  |  |  |
| State certified Nurse | 1.20 (0.20-7.18) | 0.84 |  |  |  |
| Lab technician | 3.60 (0.28 –44.82) | 0.31 |  |  |  |
| Registered nurse | 1.20 (0.14 – 10.11) | 0.86 |  |  |  |
| Nurse assistant | 2.37 (0.44-12.75) | 0.31 |  |  |  |
| **Years of experience in PMTCT(n=183)** |  |  |  |  |  |
| None | 1 |  |  |  |  |
| < 5 years | 0.40 (0.04-3.31) | 0.39 |  |  |  |
| 5 – 10 years | 0.30 (0.03-2.73) | 0.28 |  |  |  |
| >10 years | 0.16 (0.01- 2.09) | 0.16 |  |  |  |
| **Formal training on PMTCT** |  |  |  |  |  |
| Yes | 1 |  |  |  |  |
| No | 4.35 (1.44-13.09) | 0.009* | 5.03 (1.57-16.07) | **0.006\*** |  |

## Discussion

In this study, we presented the current state of PMTCT in informal health centers in the cities of Douala and Ebolowa in Cameroon. The study highlighted that almost all (99%) IHCs with ANC services integrated HIV testing in antenatal laboratory exams. This study was carried out in IHCs in Ebolowa and Douala, respectively main towns of the South and Littoral Regions of Cameroon. When we look at Cameroon national PMTCT data from 2019, we see that the coverage of HIV testing in these Regions (98.2% and 93.7% respectively) is largely above the national coverage of 82.8% **[27]**. This could lead one to think that the formal ANC services in these regions mainly include the HIV screening test in the antenatal laboratory exams package; and the informal health centers followed this trend.

Our results showed that, unlike other informal health providers that typically have little or no formal health training, the majority of those working in IHCs in our setting had received formal health training. On the one hand we have some owners of IHCs who are retired civil servants or still in service. On the other, HPs who have not been able to integrate into the formal health system. Regarding the latter, Similar results were found in studies on informal health care services in Nigeria. Indeed, in these studies, among the informal providers, there were those with adequate formal health training. They found themselves in the informal sector because they could not obtain a position in the formal healthcare system and in order to generate income and apply their knowledge **[14,28]**. On another side, it is clear from the findings of this study that most HPs have insufficient knowledge (51%) relating to PMTCT and performed worse practices (90%) towards PMTCT.

These gaps reflect the concern of some about the quality of care provided by informal providers in general **[28–30]**. However, some studies conducted in formal health facilities in Cameroon founded similar results. Those studies showed that about 40% of Health Care Workers (HCWs) had insufficient knowledge **[31–33]** and a great number (76%) had insufficient practices of PMTCT **[31]**. By this comparison with formal sector, we finally observe that in terms of poor PMTCT knowledge, and poor PMTCT practices, being informal or formal does not seem to matter since they majority of health providers in the two sectors are lacking knowledge and practices regarding PMTCT.

The item that recorded the lowest score regarding PMTCT knowledge was breastfeeding of HIV exposed child. In fact, lack of knowledge about HIV exposed infant feeding was high among our HPs. This show either lack of awareness of HPs regarding most recent WHO guidelines on HIV and infant feeding, that promote exclusive breastfeeding for the six first months of live **[34]**; or fear of vertical transmission from mother to child despite the ART coverage in the HIV mother. Same results in different settings were documented in some studies even with HCWs in formal health facilities **[35,36]**.

In this study, only 8% of HCWs would refer participants with a completed form. Indeed, the referral of the cases practiced in our context was limited to a verbal referral; consisting of a recommendation in which the provider gives just the name of the health facilities, preferably public, where the patient can ensure the continuum of care. This type of verbal and general referral is widely practiced by informal care providers **[13,14]**. In Cameroon a review of maternal deaths at Douala General hospital highlighted that all references made by IHCs were verbal without any documentation. This made it difficult for the reception hosted health facility to know prior interventions received by the patient.

Nevertheless, referrals management issue is not specific to IHCs. Even in formal health facilities, referral systems is a big issue. Even though there is a formal referral system with guidelines on referral **[37]**.

In addition, only 15% of HPs encouraged HIVPW to disclose their HIV status to their male partners and proposed them HIV counselling and screening. Yet HIV counselling and testing of male partners of pregnant women is a relevant strategy to increase the use of PTMCT services **[38–40]**.

In the multivariate logistic model, the lack of formal training in PMTCT was the only factor that was associated with both the insufficient of knowledge and practices in PMTCT. Inadequate PMTCT knowledge and practices due to lack of formal PMTCT training is not specific to IHCs. Indeed, several studies in formal health facilities have shown that the low skills of HCWs relating to PMTCT were attributable to the lack of training, which constitutes a hindrance to access to reliable and complete information **[31,41]**.

Furthermore, HPs without any experience in PMTCT were thirty-two times more at risk to have poor knowledge on PMTCT compared to those with at least 5 years’ experience. The latter used to perform PMTCT activity during a considerable period of time. Consequently, doing the same thing during a long period may lead to a better understanding and better knowledge.

These results ultimately imply the need for PMTCT programs to emphasize the training of Staff involve primarily in Mother and Child services, as training enhances knowledge retention. Also, the formative supervisions which will make it possible to better fix knowledge, should be reinforced in order to elucidate the doubts/shadow zones which the staffs encounter in their daily clinical activities, and keep them up to date with good PMTCT practices.

However, this study is likely to have a power issue due to the small sample sizes analysed.

## Conclusion

This study, HIV testing in antenatal care services was offered by the majority of IHCs. But the level of knowledge and practices related to PMTCT was very low. A good number of IHCs referred HIVPW for HIV care. Verbal referrals were the predominant referral system performed. Since IHCs receive pregnant women and perform a number of PMTCT activities, they must not be left behind in the strategy towards the achievement of elimination of HIV transmission from mother to child in Cameroon.

## Data Availability

The data that support the findings of this study are not openly available because the PhD thesis work from which this data is derived has not yet been defended.

NA

## Acknowledgements

The authors are grateful all the managers of the informal health centers who allowed us to carry out this study in their structures, all their staff who take part in this study and Cédric Djadda who helped us to collect the questionnaires in the city of Douala.

